# Integrated transcriptomics identifies key genes and pathways driving CKD to ESRD progression: implications for diagnosis and prognosis

**DOI:** 10.64898/2026.10.14.26365574

**Authors:** Xing Zhang, Yundong Xie, Huiyao Cao, Fan Ping, Honglian Yang, Kai Qu, Xia Shen

**Affiliations:** College of Pharmacy, Shaanxi University of Chinese Medicine, Xianyang, Shaanxi, China; Department of Pharmacy, the First Affiliated Hospital of Xi’an Jiaotong University, Xi’an, Shaanxi, China; Shaanxi University of Chinese Medicine Affiliated Hospital, Xianyang, Shaanxi, China; Shaanxi Provincial Hospital of Chinese Medicine, Xi’an, Shaanxi, China

**Keywords:** chronic kidney disease, end-stage renal disease, biomarkers, diagnosis, therapeutic target

## Abstract

**Background:** Chronic kidney disease (CKD) may progress to stage 5 CKD (end-stage renal disease, ESRD), but stage-associated molecular alterations remain incompletely characterized. This study aimed to identify candidate biomarkers associated with CKD and ESRD.

**Methods:** Two discovery datasets comparing CKD or ESRD with healthy controls were analyzed using differential expression analysis and weighted gene co-expression network analysis. Candidate genes were identified by intersecting differentially expressed genes with key co-expression modules, followed by protein-protein interaction network analysis using Degree, Closeness, and Maximal Clique Centrality algorithms. Candidate genes were evaluated in independent GEO validation datasets and further examined by quantitative real-time PCR and western blotting using clinical blood samples.

**Results:** Ten hub genes were identified, among which *EPB42, KLF1*, and *H3F3A* were prioritized for further evaluation. *EPB42* showed decreased expression in CKD but increased expression in ESRD, indicating an opposite-direction pattern between the two disease states. *KLF1* showed no significant difference between healthy controls and CKD but was increased in ESRD, whereas *H3F3A* was increased in CKD and further increased in ESRD. Functional enrichment analysis highlighted processes related to oxygen transport, mitochondrial function, muscle-associated processes, and inflammation, suggesting possible involvement of hypoxia-associated oxidative injury and ferroptosis-related processes.

**Conclusions:** Integrated transcriptomic and experimental analyses identified *EPB42, KLF1*, and *H3F3A* as candidate stage-associated biomarkers in CKD and ESRD. In particular, the opposite-direction expression pattern of *EPB42* may provide additional molecular information for distinguishing CKD from ESRD.

## Introduction

Chronic kidney disease (CKD) is a heterogeneous and usually progressive disorder characterized by persistent abnormalities of kidney structure or function and represents a major global health burden [1-3]. The pathogenesis and progression of CKD involve multiple molecular pathways, including the renin–angiotensin system, TGF-β/Smad signaling, and Wnt/β-catenin signaling [4, 5], as well as broader transcriptomic, proteomic, metabolomic, and other omics-related alterations [6-9]. Although therapeutic options for CKD have expanded considerably, substantial residual risk of progression to kidney failure remains [10, 11]. Early detection and accurate assessment of disease progression therefore remain important components of CKD management.

Current clinical assessment of CKD relies primarily on established indicators of kidney function and kidney damage, particularly estimated glomerular filtration rate (eGFR), serum creatinine, and urinary albumin or protein measurements [12-14]. These measures are indispensable for diagnosis, staging, and risk assessment; however, they may not fully reflect the molecular heterogeneity associated with different disease states. Recent transcriptomic, multi-omics, and metabolomic studies have demonstrated the potential of molecular signatures to provide complementary biological information and facilitate biomarker discovery in CKD [15-17]. Therefore, identifying molecular markers associated with different stages of CKD may contribute to a more detailed characterization of disease-related molecular changes.

In this study, we tried to use a novel strategy that jointly analyzes CKD and ESRD datasets, rather than examining each stage in isolation. We integrated differential expression analysis, WGCNA, and PPI network screening to identify key genes, followed by rigorous validation across four independent GEO datasets and experimental confirmation using qRT-PCR and western blotting (WB). This multi-stage, multi-cohort analytical framework allows us to capture dynamic molecular shifts during disease progression and to pinpoint biomarkers that specifically reflect the transition from CKD to ESRD. By identifying such stage-discriminatory biomarkers, our approach aims to enable earlier diagnosis, more precise staging, and optimal timing of therapeutic intervention, ultimately helping to delay or prevent the devastating progression from CKD to ESRD in clinical practice. The study design is presented in Fig 1.

**Fig 1.** Summary of research workflow. CKD: Chronic kidney disease; ESRD: End stage renal disease; DEGs: Differentially expressed genes; KM: Key module.

## Materials and methods

### Data collection and differential gene expression analysis

The gene expression matrices and corresponding sample information for GSE66494 and GSE37171 were obtained from the Gene Expression Omnibus (GEO) database. These datasets were selected as discovery cohorts because they included human samples from patients with CKD or ESRD together with healthy controls, provided accessible gene expression profiles and clinical group annotations, and contained sufficient samples for downstream differential expression and co-expression network analyses. GSE66494 comprised kidney biopsy samples from 53 patients with CKD and 8 healthy controls (HCs), whereas GSE37171 comprised peripheral blood samples from 63 patients with ESRD and 20 HCs. Detailed information on the datasets is provided in S1 Table. Expression data were normalized using the normalizeBetweenArrays function in the limma package in R (version 4.1.3). Differentially expressed genes (DEGs) were identified using the thresholds of an absolute log2 fold change (|log2FC|) ≥ 1 and an adjusted *P* value < 0.05.

### Weighted gene coexpression network analysis

In order to identify the potential key module (KM), we used the WGCNA package of R (version: 4.1.3) [18]. The network construction procedure included the following main steps: (1) clustering analysis: to screen for outliers in all the samples by clustering. (2) constructing a scale-free network: choosing β as a soft-thresholding parameter to enhance the strong correlations between genes and penalize the weak correlations, in order to ensure high scale independence and average connectivity. (3) transforming the adjacency relationship into a topological overlap matrix (TOM) and calculating the corresponding dissimilarity (1-TOM); (4) detecting modules by hierarchical clustering and using the dynamic tree cut function, using 1-TOM as the distance measure with a deepSplit value of two and a minimum size cutoff of 50 for the resulting dendrogram, and (5) identifying the KMs using the module eigengenes (MEs) to compute for the correlations between the modules and clinical information. The ME, which was the first principal component in each module, represented all the genes in each module.

### Key gene identification

To obtain overlapping genes, DEGs from the GSE66494 (DEGs1) were intersected with the KM1 and the DEGs from the GSE37171 (DEGs2) were intersected with the KM2. The overlapping genes from GSE66494 and GSE37171 were combined as the candidate key genes. The online database STRING was used to predict the PPI networks [19]. Thereafter, the Degree, Closeness, and MCC algorithms of the cytoHubba plugin for Cytoscape were used to identify candidate genes in the PPI network; the top 15 were selected as the candidate key genes [20]. The key genes that were obtained by these algorithms were intersected to identify the final hub genes.

### Function and pathway enrichment analysis

DAVID (https://davidbioinformatics.nih.gov/), which is a database for annotation, visualization, and integrated discovery [21], was used to perform Gene Ontology (GO) and Kyoto Encyclopedia of Genes and Genomes (KEGG) pathway analyses of DEGs1, DEGs2, KM1 and KM2. The ontology contained three classes, including biological process (BP), molecular function (MF), and cellular component (CC). Enrichment significance was assessed using Fisher’s exact test, and multiple-testing correction was performed using the false discovery rate (FDR). Terms with an FDR < 0.05 were considered statistically significant.

### Validation of key genes

GSE62792 and GSE142153 were obtained from the Gene Expression Omnibus (GEO) database and used as independent validation datasets for CKD and ESRD, respectively. GSE62792 included 12 patients with CKD and 6 healthy controls (HCs), whereas GSE142153 included 7 patients with ESRD and 10 HCs. Detailed information on these datasets is provided in S1 Table. Gene expression data were analyzed using the limma package, and differences in the expression of candidate genes between the disease and corresponding control groups were evaluated. A *P* value < 0.05 was considered statistically significant. Receiver operating characteristic (ROC) curve analyses were subsequently performed using the pROC package in R to assess the discriminatory performance of the candidate genes in the validation datasets. The area under the ROC curve (AUC) and the corresponding 95% confidence interval (CI) were calculated for each candidate gene.

### Samples, quantitative real-time polymerase chain reaction, and western blotting

To further validate the potential roles of the key genes in CKD and ESRD, blood samples were obtained from 10 patients with CKD at the time of diagnosis and 10 patients with ESRD. Healthy participants were recruited as controls. Informed consent was obtained from each participant, and approval of this study was granted by the Ethics Committee of Shaanxi Provincial Hospital of Traditional Chinese Medicine, Approval No: (2021) Lunshen No. (12). The pertinent information is listed in S2 Table.

Total RNA was extracted using a TRIzol Reagent (Ambion: #15596-026), and the concentration was calculated by the OD260/OD280 ratio. Thereafter, cDNA was synthesized using the HiScript II Q RT SuperMix for qPCR (+gDNA wiper) Kit, followed by further RT-qPCR (ViiA™7, Applied Biosystems). The housekeeping gene GAPDH was used as an endogenous control. The relative expressions of EPB42, KLF1, and H3F3A mRNA were calculated using the 2^−ΔΔCt^ method. S3 Table shows the primers used in this study.

The protein expression levels were confirmed by WB. Human whole blood samples were treated with a human whole blood mononuclear cell separation liquid (LDS1075, Tianjin Haoyang Biology, Tianjin, China), which contained phosphatase and protease inhibitors. The BCA Protein Detection Kit (Beyotime Biotechnology: #P0010) was used to detect the protein concentration. Subsequently, an equivalent amount of protein was separated by 10% Tris-HCl SDS-PAGE (Biofroxx: #1115GR500) and transferred onto polyvinylidene fluoride membranes (Millipore: #IPVH00010). After blocking with skimmed milk in TBST for 2 h, the membrane was further probed overnight at 4 °C using antibodies against GAPDH (Proteintech: #60004-I-Ig), EPB42 (Thermo: # PA5-10968), KLF1 (Affinity: # DF3259), and H3F3A (Affinity # DF7888). The membranes were subsequently washed with TBST, then incubated with an HRP-conjugated anti rabbit antibody at room temperature for 2 h. The membrane was immersed in enhanced chemiluminescence reagent (APPLYGEN, Beijing, China), and immunoreactive bands were assessed using a gel imaging system. Finally, Image-Pro Plus software was used to quantify the gray value of the protein bands.

### Statistical analysis

Statistical analyses were performed using IBM SPSS Statistics (version 31.0; IBM Corp., Armonk, NY, USA). Quantitative data are presented as the mean ± standard deviation (SD). For qRT-PCR analysis, ΔCt values at the biological-sample level were used for statistical comparisons, whereas normalized target protein/GAPDH ratios were used for western blotting analysis. Differences among the HC, CKD, and ESRD groups were assessed using one-way analysis of variance (ANOVA), followed by Tukey’s multiple comparison test. All tests were two-sided, and a *P* value < 0.05 was considered statistically significant.

## Results

### Identification of DEGs in the GSE66494 and GSE37171 datasets

The normalized expression data for GSE66494 and GSE37171 are shown in S1 Fig and S4 and S5 Tables. A total of 2129 DEGs1 were identified between the CKD and HC groups in GSE66494, including 679 upregulated and 1,450 downregulated genes in CKD relative to HC (S2 Fig). A total of 411 DEGs2 were identified, including 301 upregulated and 110 downregulated DEGs in ESRD relative to HC (S2 Fig).

### Weighted gene coexpression network analysis

All genes were used for the WGCNA construction. At a soft thresholding power β of 12, the coexpression network met the scale-free topology threshold (R^2^ = 0.85), and the mean connectivity was close to 0 in the two datasets. We identified 25 modules in the GSE66494 by WGCNA (Fig 2A). Based on the Spearman correlation coefficient, a heat map of the module–trait relationship was drawn to assess the relationships between modules (Fig 2C). The modules “salmon” and “pink” were highly associated with CKD; on the eigengene dendrogram, the salmon module had superior correlation with the CKD status (Fig 2E). Therefore, the salmon module (r = 0.81, *P* = 1e-14) was chosen as CKD-related KM (Fig 2G). In the GSE37171, 10 modules were discovered, and the module “turquoise” (r = 0.80, *P* = 2e-26) showing highly positive link with ESRD (Fig 2B, 2D). The turquoise module showed a strong positive correlation with ESRD status and was therefore selected as the ESRD-related key module (Fig 2D). The eigengene dendrogram is shown in Fig 2F, and module membership was strongly correlated with gene significance in the turquoise module (Fig 2H).

**Fig 2.** WGCNA and Significant modules associated with CKD and ESRD. The cluster dendrograms of the coexpressed genes in (A) CKD and (B) ESRD are shown, with each module represented by a different color. Heatmaps of the module–trait relationships in (C) CKD and (D) ESRD are shown. Hierarchical clustering dendrograms of (E) CKD and MEs and (F) ERSD and MEs are shown. Correlation charts between the gene members of the (G) salmon and (H) turquoise modules, as well as the gene significance, are shown. The *P* values < 0.05 indicating statistical significance.

#### Key gene identification

On the Venn diagram, 70 CKD genes were all present in DEGs1 and salmon modules (Fig 3A), and 111 ESRD genes from the DEGs2 and turquoise modules overlapped (Fig 3B). After combining the genes, we obtained 181 key gene candidates for CKD and ESRD. The top 15 genes in the PPI network were selected and are shown in S6 Table, the intersection of these genes was used to construct a Venn diagram, resulting in 10 key genes, including *EPB42, KLF1, H3F3A, RPS11, ANK1, PRMT6, EP300, GLTSCR2, RXRB*, and *MYSM1* (Fig 3C).

**Fig 3.** Identification of the key genes in CKD and ESRD. Venn diagrams of the DEGs in the datasets of (A) CKD and (B) ESRD and (C) the key genes are shown.

### Function and pathway enrichment analyses of DEGs

The GO and KEGG enrichment results for DEGs1 and DEGs2 are shown in Fig 4A–4E and S7 and S8 Tables. GO enrichment analysis indicated that DEGs1 and DEGs2 were associated with several oxygen transport- and hemoglobin-related terms, including oxygen transport activity, oxygen binding, hemoglobin complex, haptoglobin binding, and heme binding. Mitochondrial- and metabolic-related terms, including mitochondrion, ATP binding, metal ion binding, and succinyl-CoA catabolic process, were also enriched. In addition, DEGs1 were enriched in several muscle-related processes and structures, such as muscle contraction and sarcomere, as well as estrogen metabolic process and calcium ion binding.

**Fig 4.** Function and pathway enrichment analysis of CKD and ESRD gene. The top 10 GO terms from the CKD dataset in the (A) BP, CC, and MF classes are shown. The top 10 GO terms from the ESRD dataset in the (B) BP, CC, and MF classes are shown. The GO function enrichment analysis of the salmon (C) and turquoise (D) modules are shown. (E) The KEGG pathway enrichment analyses of the CKD (DEGs1, DEGs1_down, and DEGs1_up) and ESRD (DEGs2, DEGs2_down, and DEGs2_up) datasets and the salmon and turquoise modules are shown. The outermost layer of the plot shows Gene Ontology (GO) ID numbers. The dots represent the logFC values, with red dots indicating upregulated genes and blue dots indicating downregulated genes. The higher the innermost bar, the greater the enrichment score (A-B). The “count” represents the number of genes assigned to the corresponding KEGG pathway; the color indicates significance, with a bluer shade representing greater significance (C-E). logFC: log fold change; GO: Gene Ontology; KEGG: Kyoto Encyclopedia of Genes and Genomes.

KEGG enrichment analysis showed that DEGs1 were mainly enriched in pancreatic secretion, the MAPK signaling pathway, vascular smooth muscle contraction, and thyroid hormone secretion, whereas DEGs2 were enriched in the FoxO signaling pathway and nucleotide metabolism. Further examination of the expression direction of genes mapped to these pathways showed that genes associated with pancreatic secretion, the MAPK signaling pathway, and vascular smooth muscle contraction were predominantly downregulated in DEGs1. In DEGs2, genes involved in nucleotide metabolism were predominantly upregulated, whereas those mapped to the FoxO signaling pathway were predominantly downregulated (Fig 4E).

### Function and pathway enrichment analysis of key module

To further characterize the biological functions associated with the key modules, GO and KEGG enrichment analyses were performed. A total of 39 GO terms, including 19 BP, 11 CC, and 9 MF terms, together with two KEGG pathways, were enriched in the salmon module. Up to the top 10 enriched GO terms in each category are shown in Fig 4C. BP terms were mainly associated with Ras- and Rap-related signal transduction, whereas MF terms primarily reflected enzymatic activities related to these signaling processes. CC terms were mainly associated with the mitochondrial inner membrane, cytoplasm, and nucleus. KEGG pathway analysis identified nucleocytoplasmic transport and lysosome as enriched pathways (Fig 4E).

For the turquoise module, 432 GO terms were enriched, including 302 BP, 69 CC, and 61 MF terms, together with 32 KEGG pathways. The top 10 enriched GO terms in each category are shown in Fig 4D. BP terms were predominantly associated with transcription from the RNA polymerase II promoter, whereas MF terms were mainly related to corresponding transcription-associated molecular functions. CC terms were primarily associated with the extracellular matrix and chromatin. KEGG analysis further showed enrichment of several signaling pathways, including the calcium, Rap1, and Ras signaling pathways (Fig 4E).

Notably, Ras- and Rap-related signaling was represented in the enrichment results of both the salmon and turquoise modules. Overall, the enrichment profiles of the two key modules showed functional overlap in signaling-related processes, while also revealing module-specific enrichment patterns.

### Validation of key genes using other datasets

In the GSE62792 validation cohort, *EPB42* showed a significant difference in expression between patients with CKD and healthy controls (HCs), whereas *KLF1* did not show a statistically significant difference (Fig 5A). ROC analysis showed AUCs of 1.000 (95% CI: 1.000–1.000) for *EPB42* and 0.806 (95% CI: 0.597–1.000) for *KLF1*, suggesting potential discriminatory value in this validation cohort (Fig 5B). *MYSM1* was not detected in the GSE62792 dataset. In the GSE142153 validation cohort, *EPB42* and *H3F3A* showed significant differences in expression between patients with ESRD and HCs (Fig 5C). ROC analysis yielded AUCs of 1.000 (95% CI: 0.836– 1.000) for *EPB42* and 0.806 (95% CI: 0.800–1.000) for *H3F3A*, respectively (Fig 5F).

**Fig 5.** Key gene expression and diagnostic value in CKD and ESRD. (A, C) Comparisons of the changes in the expressions of key genes in the GSE62792 (CKD) and GSE142153 (ESRD) datasets.(B, F) Receiver Operating Characteristic (ROC) curves of the 10 key genes in the GSE62792 and GSE142153 validation datasets, respectively, with area under the curve (AUC) values indicating diagnostic value.(D, E) Comparisons of the changes in the expressions of *EPB42, KLF1*, and *H3F3A* in the GSE62792 and GSE142153 datasets. The *P* value < 0.05 indicating statistical significance. ROC: Receiver Operating Characteristic; AUC: Area under the curve.

Subsequently, the expression patterns of *EPB42, KLF1*, and *H3F3A* were examined across the four datasets (Fig 5D and 5E). *EPB42* showed lower expression in CKD datasets but higher expression in ESRD datasets relative to their corresponding HCs, indicating an opposite-direction expression pattern between the two disease states. *KLF1* showed no significant difference between CKD and HC groups but tended to show higher expression in ESRD. *H3F3A* generally showed higher expression in both CKD and ESRD datasets relative to their corresponding control groups.

### Samples, quantitative real-time polymerase chain reaction, and western blotting

To validate the expression levels of *EPB42, KLF1*, and *H3F3A*, peripheral blood samples from patients with CKD, patients with ESRD, and HCs were analyzed by qRT-PCR and western blotting (Fig 6). qRT-PCR analysis showed that *EPB42* was significantly downregulated in CKD but upregulated in ESRD. *KLF1* showed no significant difference between CKD and HCs but was significantly upregulated in ESRD, whereas *H3F3A* was significantly upregulated in both CKD and ESRD. Western blotting showed corresponding changes in *EPB42, KLF1*, and *H3F3A* protein expression, consistent with the qRT-PCR findings and bioinformatics-derived predictions.

**Fig 6.** The expressions of EPB42, KLF1, and H3F3A were validated by quantitative real-time PCR (qRT-PCR) and Western blotting (WB). (A–C) Expression of genes at the mRNA level by qRT-PCR. (D) Expression of genes at the protein level by WB. The main figure for presentation purposes and that the corresponding full-length original blot images are provided in S1 File. qRT-PCR, quantitative real-time PCR; WB, Western blotting. \**P*< 0.05; \*\**P*< 0.01; \*\*\**P*< 0.001. ns, no significance.

## Discussion

The present study integrated differential expression analysis, WGCNA, PPI network analysis, external transcriptomic validation, and experimental validation to identify molecular features associated with CKD and ESRD; ten hub genes were initially identified, among which *EPB42, KLF1*, and *H3F3A* showed relatively consistent expression patterns across different validation analyses and exhibited distinct stage-associated profiles, with *EPB42* showing an opposite-direction expression pattern (lower in CKD and higher in ESRD), *KLF*1 showing no significant change in CKD but an increase in ESRD, and *H3F3A* being increased in CKD and further increased in ESRD. Although conventional clinical indicators such as eGFR, serum creatinine, and urinary protein remain important for CKD diagnosis and staging, these three genes may provide additional information on biological changes associated with different disease states. Notably, the opposite-direction expression pattern of *EPB42* represents a distinctive finding of this study and, together with the different expression characteristics of *KLF1* and *H3F3A*, suggests their potential value as stage-associated molecular markers for distinguishing CKD from ESRD and for providing molecular information beyond conventional clinical indicators [22, 23]. Thus, the main innovation of this study lies in its integrated multi-omics and experimental validation strategy and in identifying the stage-associated expression patterns of *EPB42, KLF1*, and *H3F3A* as candidate molecular features that may contribute to differentiating CKD from ESRD and complement conventional clinical indicators.

*EPB42* encodes erythrocyte membrane protein 4.2 [24], an important component of the erythrocyte membrane skeleton that contributes to membrane stability, deformability, and erythrocyte integrity [25]. Previous studies have shown that alterations in erythrocyte membrane structure and deformability are closely associated with erythrocyte survival and hematological homeostasis in CKD. In the present study, *EPB42* exhibited a distinctive opposite-direction expression pattern, with decreased expression in CKD but increased expression in ESRD, distinguishing it from the other candidate genes identified. This finding, together with the enrichment of oxygen transport- and hemoglobin-related biological processes, suggests that *EPB42* may be closely associated with stage-related changes in erythrocyte function and oxygen-carrying capacity [26-28]. As illustrated in Fig 7, reductions in erythrocyte number and hemoglobin concentration may impair systemic oxygen delivery, while disturbances in renal oxygen utilization may further contribute to an imbalance between oxygen supply and demand [29]. Persistent hypoxic stress may subsequently affect mitochondrial function and cellular energy metabolism [30], whereas mitochondrial dysfunction, uremic toxin accumulation, and disturbances in nucleotide and glutathione metabolism may further aggravate metabolic and oxidative stress [31-34]. These interconnected processes provide a plausible biological framework linking EPB42-associated erythrocyte changes with oxygen homeostasis, mitochondrial dysfunction, and oxidative injury during CKD and ESRD. Moreover, the involvement of glutathione metabolism and mitochondrial dysfunction raises the possibility that ferroptosis-related processes may participate in these molecular alterations [35-37]. Taken together, the bidirectional expression pattern of *EPB42*, its established role in erythrocyte biology, and its close association with oxygen transport-related pathways highlight *EPB42* as a distinctive stage-associated biomarker with potential value for differentiating CKD from ESRD and for improving the molecular characterization of disease-stage transitions.

**Fig 7.** Factors in the transition from CKD to ESRD.

*KLF1* and *H3F3A* exhibited distinct expression patterns and may reflect different biological processes associated with CKD and ESRD. *KLF1* is a key transcriptional regulator of erythroid differentiation and controls the expression of multiple genes involved in erythrocyte development and function [38]. In the present study, *KLF1* showed no significant alteration in CKD but was increased in ESRD, suggesting a closer association with the advanced disease state. Given its central role in erythropoiesis, changes in *KLF1* expression may be related to alterations in erythrocyte production, hemoglobin homeostasis, and oxygen delivery during advanced kidney dysfunction. Together with the oxygen transport-related enrichment observed in this study, these findings suggest a possible link between *KLF1* dysregulation, impaired oxygen homeostasis, and hypoxia-associated oxidative stress. Disturbances in erythropoiesis and redox balance may also influence iron homeostasis and cellular susceptibility to ferroptosis-related injury [39]. In contrast, *H3F3A* encodes histone H3.3, a histone variant involved in chromatin organization, nucleosome formation, and transcriptional regulation [40]. *H3F3A* expression was increased in CKD and remained elevated in ESRD, indicating a broader association with disease-related molecular alterations.

Histones and nucleosomes can be released during neutrophil extracellular trap (NET) formation, while NETosis and chronic inflammatory responses have been implicated in CKD-related tissue injury [41-43]. Increased inflammatory activity and excessive NET formation may enhance oxidative stress and lipid peroxidation [44], providing a potential connection between H3F3A-associated chromatin changes, inflammation, and ferroptosis-related processes. Taken together, the distinct expression characteristics of *KLF1* and *H3F3A* suggest that they may represent complementary molecular features of CKD and ESRD, with *KLF1* being more closely linked to erythroid and oxygen-transport regulation and *H3F3A* potentially reflecting chromatin- and inflammation-associated alterations. These findings provide additional biological support for their potential use as stage-associated molecular biomarkers and offer mechanistic clues for understanding the molecular changes accompanying advanced kidney disease.

From a clinical perspective, the distinct expression patterns of *EPB42, KLF1*, and *H3F3A* may be useful for distinguishing different disease states. *EPB42* showed opposite expression directions in CKD and ESRD, *KLF1* was mainly increased in ESRD, and *H3F3A* showed a sustained increase across CKD and ESRD. These complementary expression patterns suggest that combining markers with different biological characteristics may provide more information than relying on a single marker. Such molecular markers may complement conventional measures of kidney function and damage and contribute to more refined disease staging and earlier recognition of molecular changes associated with advanced kidney disease [45]. In addition, *EPB42, KLF1*, and *H3F3A* may provide candidate molecules for future mechanistic and pharmacological studies, thereby offering new directions for understanding the biological changes associated with CKD and ESRD.

Despite the encouraging findings obtained in this study, several factors should be considered when interpreting the current results. The discovery datasets were selected to provide complementary, hypothesis-generating molecular information from different disease stages and tissue compartments, rather than to serve as interchangeable replicates of the same tissue. Specifically, GSE66494 is based on kidney biopsy samples from patients with CKD, whereas GSE37171 is based on peripheral blood samples from patients with ESRD. This design allowed us to explore disease-associated molecular features in both renal tissue and the systemic circulation. However, because tissue origin and cellular composition can substantially influence gene expression profiles, the two datasets should not be directly compared as if they represented the same biological context, and the observed transcriptional variation may partly reflect tissue-specific or cell-composition-related effects. Therefore, overlapping or consistent candidate signals should be interpreted as exploratory and hypothesis-generating, and validation in tissue-matched and larger cohorts is warranted. In addition, ROC analyses were conducted in independent validation cohorts, but the relatively small sample sizes may result in optimistic or unstable estimates of discriminatory performance, particularly for biomarkers with very high AUC values. Therefore, these ROC findings should be interpreted as preliminary evidence of discriminatory potential, and further validation in larger independent cohorts will be needed to confirm their stability and generalizability.

## Conclusions

In conclusion, this study used an integrative dual-cohort transcriptomic strategy that combined differential expression analysis, WGCNA, PPI network analysis, external transcriptomic validation, and experimental validation in clinical blood samples. We first identified ten hub genes, then prioritized EPB42, KLF1, and H3F3A because their expression patterns were consistent across validation analyses. These genes showed distinct stage-related changes: *EPB42* decreased in CKD but increased in ESRD; *KLF1* showed no significant change in CKD but increased in ESRD; and *H3F3A* increased in CKD and further increased in ESRD. These patterns suggest that the three genes may help distinguish CKD from ESRD and reflect different molecular stages. Functional enrichment implicated oxygen transport, mitochondrial function, muscle-related processes, and inflammation, suggesting possible roles for hypoxia-related oxidative injury and ferroptosis. The main innovation is the dual-cohort design, which goes beyond single-disease comparisons, and the identification of *EPB42* as a first-in-class bidirectional candidate for monitoring the CKD-to-ESRD transition. Overall, these findings provide a transcriptomic basis for future non-invasive diagnostic and targeted therapeutic strategies and support further validation of *EPB42, KLF1*, and *H3F3A* in larger clinical cohorts.

## Data Availability

These data were derived from the following resources available in the public domain: Gene Expression Omnibus (GEO) database http://www.ncbi.nlm.nih.gov/geo/.

## Supporting information

**S1 Table. The datasets detail information from GEO**. CKD indicates chronic kidney disease; HC indicates healthy control

**S2 Table. Clinical information of samples enrolled from Shaanxi Provincial Hospital of Traditional Chinese Medicine**.

**S3 Table. Oligo sequences used in quantitative real-time PCR**.

**S1 Fig. GEO dataset preprocessing**. GEO indicates Gene Expression Omnibus. A, B. Gene expression level of the GSE66494 (A) and GSE37171 (B) datasets before preprocessing. C, D. Gene expression level of the GSE66494 (C) and GSE37171 (D) datasets after preprocessing. The x-axis is the sample, and its corresponding detail label is shown in the Table S4, 5.

**S4 Table. The sample detail label from GSE66494**.

**S5 Table. The sample detail label from GSE37171**.

**S2 Fig. Identification of DEGs in the GSE37171 and GSE66494 datasets**. A. The volcano map of GSE66494 dataset. B. The volcano map of GSE37171 dataset. Blue represents down-regulated genes, red represents up-regulated genes, and black represents genes with no significant difference in expression.

**S6 Table. The top 15 genes by Degree, Closeness, and MCC algorithms**.

**S7 Table. The top 10 GO term from CKD of BP, CC, MF**.

**S8 Table. The top 10 GO term from ESRD of BP, CC, MF**.

**S1 File. Raw qRT-PCR data, western blot densitometry data, and uncropped blot images**.

## Notes

### Competing Interest Statement

The authors have declared no competing interest.

### Author Declarations

Informed consent was obtained from each participant, and approval of this study was granted by the Ethics Committee of Shaanxi Provincial Hospital of Traditional Chinese Medicine, Approval No: (2021) Lunshen No. (12).

